# Birth history is associated with whole-blood and T-cell methylation patterns in relapse onset multiple sclerosis

**DOI:** 10.1101/2022.03.24.22272917

**Authors:** Maria Pia Campagna, Alexandre Xavier, Jim Stankovich, Vicki Maltby, Mark Slee, Trevor Kilpatrick, Rodney J Scott, Helmut Butzkueven, Jeannette Lechner-Scott, Rodney Lea, Vilija Jokubaitis

## Abstract

**Background:** Pregnancy in women with multiple sclerosis (MS) is associated with a reduction of long-term disability progression. The mechanism that drives this effect is unknown, but converging evidence suggests a role for epigenetic mechanisms altering immune and/or central nervous system function.

**Objectives:** We aimed to identify whole blood and immune cell-specific DNA methylation patterns associated with parity in relapse-onset multiple sclerosis.

**Methods:** We compared whole-blood methylation patterns between 96 matched pairs of nulligravida and parous females with MS (n=192). Parity was defined as at least one term or pre-term birth, and nulligravida was defined as no prior pregnancies. Methylation was measured with Illumina EPIC arrays, and data was pre-processed and statistically analysed using the *ChAMP* package. Cell-type proportions were estimated using the *EpiDISH* package, and cell-specific analysis conducted using linear regression. Gene-set enrichment analysis (GSEA) was performed with ToppGene API and GOmeth. Methylation age was calculated with the *methyAge* package. Methylation age acceleration (MAA) was calculated by regressing methylation age on chronological age. FDR<0.05 was used to assess significance.

**Results:** The median time from last pregnancy to blood collection was 16.66 years (range = 1.45 – 44.42 years). We identified 903 differentially methylated positions (DMPs) in whole blood; 365 were hypomethylated and 528 were hypermethylated in parous women. We further identified two differentially methylated regions (DMRs) in *CRYGN* on Chromosome 7 and an intergenic region on Chromosome 15. There were four and eight cell type specific DMPs in CD4+ and CD8+ cells, respectively. Differentially methylated genes were enriched in neuronal plasticity pathways. Parity was associated with reduced MAA by a mean of 1.44 to 2.27 years using the PhenoAge (p = 0.002) and GrimAge (p = 0.005) algorithms.

**Conclusion:** Whole-blood methylation patterns are associated with birth history in females with relapse-onset multiple sclerosis. We found enrichment of differentially methylated genes encoding neuronal processes and reduced MAA in parous women. These methylation changes could mediate the long-term benefit of pregnancy for disease progression in multiple sclerosis.

## Introduction

Multiple sclerosis (MS) is most prevalent in females, with a sex ratio of 3:1^1^. It is frequently diagnosed between 20-40 years of age, the prime reproductive years for women. Understanding the effect of pregnancy on disease activity and progression is a priority for women with MS (wwMS) and their care teams.

Pregnancy has been shown to reduce MS relapse rates and short- and long-term disability outcomes in wwMS, regardless of the outcome of the pregnancy^2^. With respect to long-term outcomes, in a study of 2,557 wwMS, a history of childbirth delayed the onset of a clinically isolated syndrome (CIS, the first demyelinating event indicative of a future MS diagnosis) by 3.4 years^3^. Additionally, in the largest real-world study of 1830 wwMS, one or more pregnancies after MS onset were associated with lower disability scores after ten years^4^. Notably, the protective effect of pregnancy in this cohort was four-fold greater than that of first line DMT exposure in the same timeframe^4^. The biological mechanisms underpinning these long-term effects of pregnancy are not understood. As the effect of pregnancy on age at CIS onset^3^ and disability progression extends for years beyond birth^4^, it cannot be explained exclusively by transient hormonal and immunological changes during pregnancy.

Epigenetic mechanisms regulate gene expression in a dynamic and reversible manner. DNA methylation is a key epigenetic mechanism. The presence or absence of a methyl group on cytosine-phosphate-guanine (CpG) dinucleotides generally activates or represses gene transcription, respectively. Epigenetic mechanisms are influenced by life events and environmental factors, including the multitude of physiological and hormonal changes of pregnancy. DNA methylation enzymes are specifically influenced by estrogen signalling, which increases in pregnancy and peaks in the third trimester. Converging evidence outlines a role for DNA methylation in the effect of pregnancy on outcomes in wwMS through altering immune and central nervous system (CNS) function: 1) estrogen signalling influences DNA methylation enzymes^5^, 2) pregnancy has been shown to reduce immune epigenetic age in women without MS^6^, and 3) pregnancy induces changes in the expression of immune-activation^7^ and axon-guidance^8^ genes in wwMS for up to 19 years after pregnancy. However, no epigenome-wide association study (EWAS) of parity in wwMS has been reported to date.

The objective of this study was to understand the long-term impact of parity on DNA methylation patterns in women with relapse-onset multiple sclerosis. We first sought to identify whole-blood and immune cell-specific DNA methylation patterns, across autosomes, associated with parity. Secondly, we aimed to compare methylation age acceleration (MAA) between nulligravida and parous wwMS, to determine whether reductions in MAA reported in health were also evident in an MS cohort.

## Materials and Methods

### Ethics approvals

Ethics approval for the collection of demographic, clinical, treatment and pregnancy history data via the MSBase Registry^9^ was obtained from the Alfred Health Human Research Ethics Committee (528/12), and institutional review boards at all participating centres. Approval for the collection of genetic data was obtained from the Australian National Mutual Acceptance Scheme (HREC/13/MH/189). Written informed consent was obtained from participants as per local laws at each study site.

### Clinical data collection

This study utilised clinical data from the MSBase Registry, an international, prospective, observational MS clinical outcomes register. Data are collected in a unified manner, and include patient demographics, expanded disability status scale (EDSS) scores, relapse, treatment and pregnancy data, as previously described^9,10^.

### Participant recruitment, parity definitions and sample collection

Whole-blood samples were obtained from 1,984 participants. From this cohort, we selected 192 matched participants based on geographical location (Australia), sex (female), birth history availability (nulligravida or parous) and age (groups age-matched within three years, **Supplementary Fig. 1**).

DNA methylation is associated with age^11^ and geographical location^11^. Therefore, we restricted participants to Australians matched by age (within three years). Participants were also matched by Age-Related Multiple Sclerosis Severity (ARMSS) scores^12^ due to non-negligible differences between nulligravida and parous groups (**Table 1**). Participants were matched using the *optmatch* package^13^ in the R statistical environment.

**Table 1.**
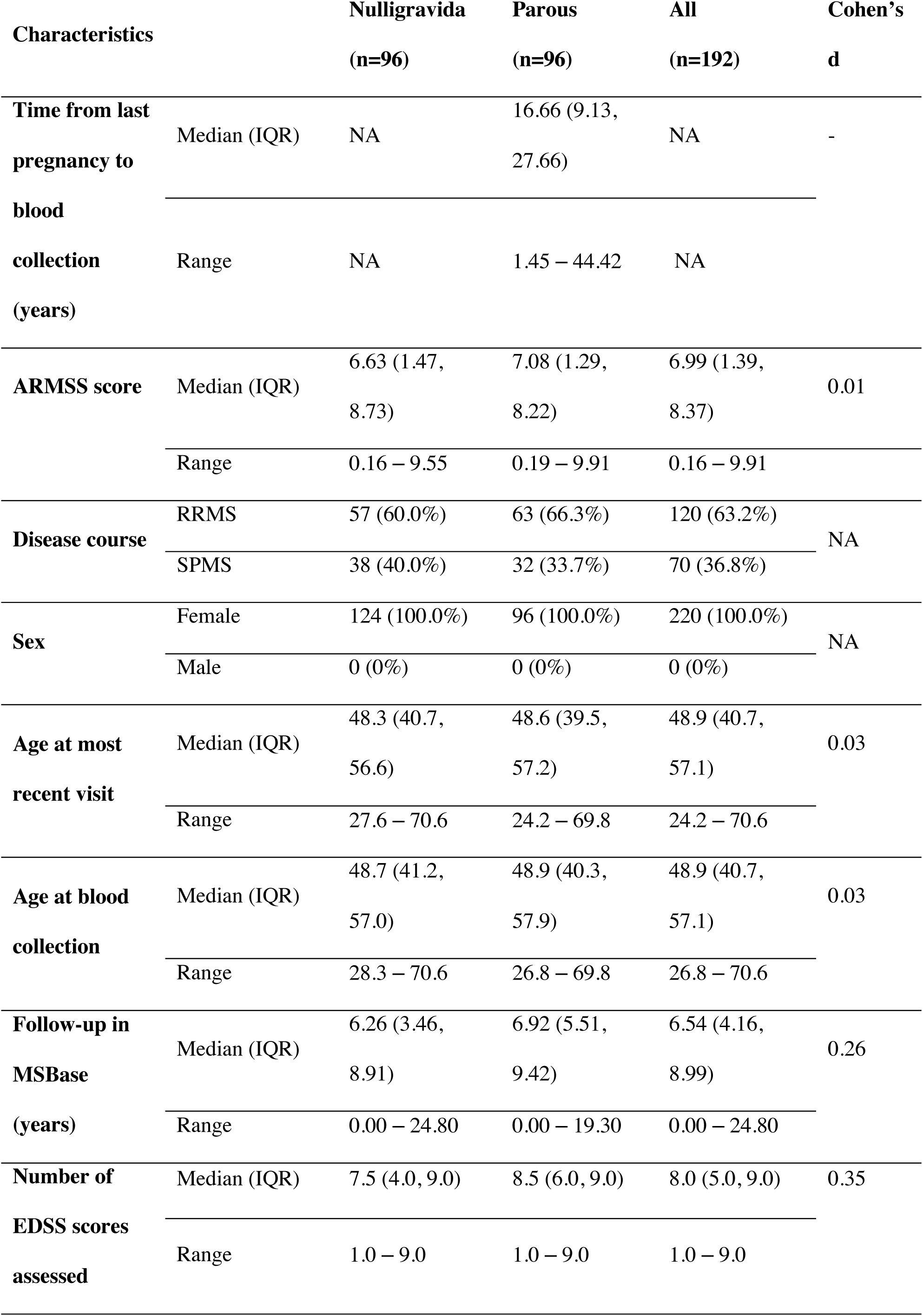
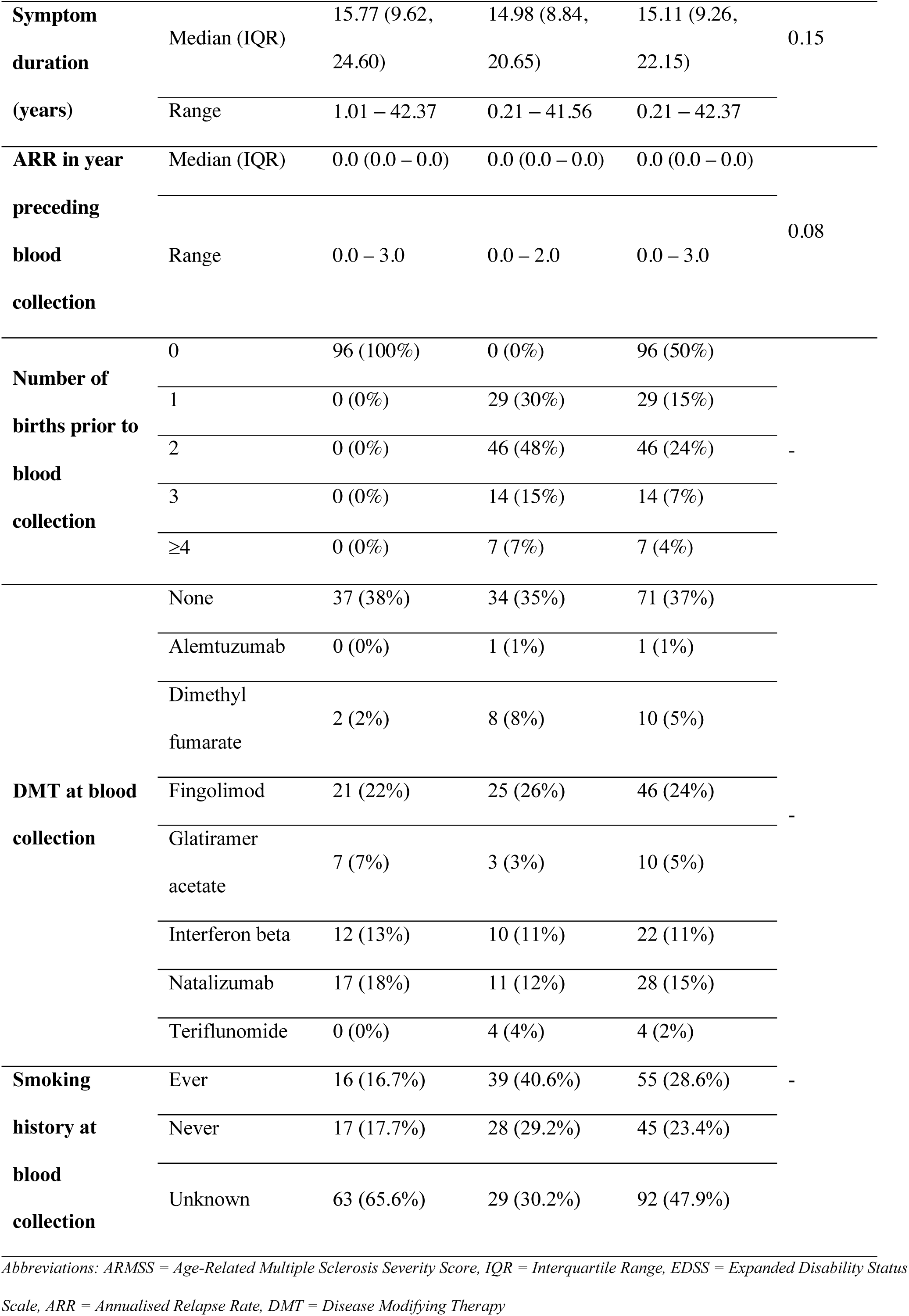
Cohort summary statistics.

The timing of pregnancy effects on methylation patterns remains unclear in wwMS, as does the impact of pregnancies resulting in miscarriage or termination compared to birth. We therefore excluded gravida women (i.e., those experiencing a miscarriage or induced abortion only) and restricted study inclusion to women who had at least one preterm or term birth prior to the date of blood collection, or those who were nulligravida. We included wwMS from the Royal Melbourne Hospital (VIC, n=73), Box Hill Hospital (VIC, n=56), John Hunter Hospital (NSW, n=25), and Flinders Medical Centre (SA, n=38). A total of 96 nulligravida and 96 parous females with RMS were included in this study (n=192).

### DNA extraction

At each site, genomic DNA was extracted from whole blood using standard protocols and procedures.

### Methylation arrays

DNA samples were processed for methylation arrays at the Hunter Medical Research Institute (NSW). DNA quantity and quality were assessed using Qbit (Invitrogen™, USA) and TapeStation (Agilent™, USA), respectively. Samples meeting concentration and quality requirements were bisulfite converted using the EZ-DNA Methylation™ Kit (Zymo) according to manufacturer guidelines. Converted DNA was hybridised to Illumina Methylation EPIC BeadChip arrays (EPIC arrays). Samples were randomised based on clinic site using the *OSAT* R package to avoid batch effects. EPIC arrays were read using an iScan (Illumina™) and raw Idat files were produced for analysis.

### Genotyping arrays

Genomic DNA was sent from participating study sites to the Center for Genome Technology, John P. Hussman Institute for Human Genomics, University of Miami, for quality assessment and genotyping. Genotyping was performed in two batches using Illumina Multi-ethnic genotyping array (MEGA^EX^) arrays. Genotype calling was conducted in GenomeStudio v2.0 (Illumina).

### DNA methylation analysis pipeline

Our EWAS analysis was informed by the guidelines described in Campagna et al. (2021)^14^. The *Chip Analysis Methylation Pipeline (ChAMP)* Bioconductor package^15^ was used for methylation data pre-processing in the R statistical environment. Raw Idat files were filtered to exclude low quality samples (failed to successful probe ratio > 0.1), low quality probes (detection p-value > 0.01, bead count < 3 in ≥5% of samples), non-CpG probes, SNP-related probes, non-autosomal probes, and multi-hit probes. Additional multi-hit probes were excluded based on Pidsley (2016) Supplementary Table 1^16^. Beta values were normalised using the beta-mixture quantile (BMIQ) method^17^. Batch effects at the array and chip level were identified with singular value decomposition (SVD) analysis^18^, and corrected for using the *Combat* algorithm^19^.

### Primary differential methylation analysis

Differential methylation (Δ_meth_) between nulligravida and parous groups was identified at the single CpG level i.e. differentially methylated positions (DMPs), and genomic region level i.e. differentially methylated regions (DMRs) using the filtered and normalised beta matrix, as previously described^14^. We used the *ChAMP* function *champ*.*DMP* to implement an unadjusted logistic model of methylation level at each probe and parity group. A false discovery rate (FDR) threshold of 0.05 was used to assess statistical significance for all analyses. Methylation beta values equate to percentage methylation, and thus going forward we report methylation differences (effect size) as a percentage (e.g., Δ_meth_ of 0.01 = 1%). DMPs with an Δ_meth_ less than 1% were removed to avoid false positives produced from technical error.

We identified DMRs using a two-pronged approach. Firstly, with the *DMRcate* R package^20^ using the following parameters: at least three DMPs within 1000bp of the adjacent DMP, a DMP and DMR threshold of FDR < 0.05. Secondly, using the DMP list to identify at least three DMPs with an FDR < 0.05 and the same direction of effect, located within 1000bp of each other. The validity of this strategy to identify DMRs in studies with small sample and/or effect sizes has previously been shown^21,22^.

As methylation can be cell type specific, immune cell type proportions were estimated to confirm that differential methylation in whole blood was not driven by differences in cell type proportions. Immune cell type proportions were estimated using the *EpiDISH* R package^23^, using methylation M-values and the reference-based *CIBERSORT* algorithm^24^. Subsequently, cell-type specific DMPs (csDMPs) were identified using a modified version of the cellDMC function of the *EpiDISH* R package^23^. We used linear regression where the outcome was methylation M-value and the predictors were cell type proportion estimate, and an interaction term of cell type proportion and parity. A separate model was run for each cell type. We used a genome-wide threshold of p ≤ 9×10^−8^ to assess statistical significance.

### Sensitivity analyses

Sensitivity analyses were performed to assess the potential impact of a series of demographic, clinical, biological, and environmental covariates on the primary methylation analysis. Covariates were selected based on non-negligible differences between groups (Cohen’s d > 0.15), or *a priori* selected. They included symptom duration, annualised relapse rate (ARR), cell type proportion estimates (B cells, CD4+ cells, CD8+ cells, NK cells, monocytes, and granulocytes), and methylation age acceleration (PhenoAge and GrimAge). Environmental factors including treatment at blood collection (yes or no) and smoking status at blood collection (ever or never) were also tested. An FDR threshold of 0.05 was used to assess statistical significance for all sensitivity analyses.

Nulligravida and parous participants were matched by age at blood collection and ARMSS scores (n=192, 96 pairs) and the difference in methylation at each probe (Δ_meth_) was calculated. Subsequently, the correlation between each Δ_meth_ and covariate was tested. Pearson’s correlation tests were used for continuous covariates (ARR, symptom duration, years follow-up in MSBase and number of available EDSS scores available). ANOVA tests were used for categorical covariates (treatment and smoking status). For categorical variables, pairs were required to have the same value for the correlation with methylation to be tested. Of 96 pairs in total, 40 pairs were on treatment at blood collection and 14 were off treatment, while eight pairs were ‘ever’ smokers at blood collection and two were ‘never’ smokers. Smoking history was unavailable for the remaining pairs. DMPs were filtered for 2,622 known smoking-associated CpGs identified by Johanes et al. (2016, Supplementary Table 2)^25^ due to the known effect of smoking on the methylome, and limited smoking data available for this cohort.

### Single Nucleotide Variant analysis

Quality control was performed with *PLINKv1*.*9*^26^. Single Nucleotide Variants (SNVs) were excluded based on low call rate (<95%), low minor allele frequency (MAF < 0.05), violation of Hardy–Weinberg equilibrium (p < 1×10^−5^), monomorphism and non-autosomal location. Samples were excluded based on sex inconsistencies, low call rate (<95%) and relatedness (pi-hat > 0.05). Relatedness was assessed using Identity by Descent (IBD) analysis in *PLINKv1*.*9*, followed by confirmation in *KING*^27^. Principal components (PC) analysis was implemented in *EIGENSTRAT*^28^. PCs were projected to 1000 Genomes Project^29^ data to assess population stratification effects, and exclude population outliers. Genotypes were then imputed using Haplotype Reference Consortium^30^ on the Michigan Imputation Server (https://imputationserver.sph.umich.edu/index.html#!), and converted to genotype calls in *PLINKv1*.*9*.

### Methylation quantitative trait loci (mQTL) analysis

Differential methylation at certain genetic loci may be influenced by the underlying SNVs at or near that site, known as methylation quantitative trait loci (mQTLs). Therefore, we tested the relationship between genotype and methylation at CpGs in the identified DMR to determine whether differential methylation was associated with, or independent of, underlying genotype.

We extracted genotypes at SNVs located five kilobases (kb) up and downstream of DMR^Chr21^ boundaries using the *KRIS* R package^31^, and assessed linkage disequilibrium (LD) using bivariate correlations of genotype frequencies with a significance threshold of p < 0.05.

To test if differential methylation within DMRs was driven by genetic effects rather than parity, we performed a linear regression with methylation as the dependent variable and genotype and parity as the independent variables.

### Multi-factor feature selection

Elastic net regression is a form of penalised regression that is useful for uncovering multiple conjoint effects in datasets with correlated features (e.g., methylation) and a greater number of features than samples (p>>>n). This method can be useful for identifying important features with greater sensitivity than conventional EWAS analyses. We used machine learning to build an elastic net regression model to identify CpGs at which methylation was associated with parity, inputting beta values at approximately 748,000 CpGs. Samples were split into training (n=134) and testing sets (n=58) to reduce overfitting. The model was trained using a cross-validation resampling method with 10 iterations, with the train function of the *caret* R package^32^. The optimal alpha value was used in a subsequent k-fold cross-validation elastic net regression to identify the minimum lambda value; using the *cv*.*glmnet* function of the *glmnet* R package^33^. These alpha and lambda values were used in the final elastic net regression model that was applied to the testing set using the glmnet function of *glmnet* R package^33^. Features (CpGs) identified by the model to be associated with parity were compared to DMPs and DMRs identified in the primary analysis, as well as mapped to genes for GSEA performed as described above.

### Gene-set enrichment analysis (GSEA)

We used gene-set enrichment analysis (GSEA) to generate hypotheses about the functional consequence of differentially methylated genes between nulligravida and parous women. All CpGs that were associated with parity in the primary differential methylation analysis and elastic net regression were used as input. We conducted GSEA using two methods. Firstly, the *ToppGene* online application programming interface (API)^34^ which takes an FDR ranked gene list ranked as input, with hypomethylated and hypermethylated genes analysed separately. Secondly, we used the *GOmeth* function^35^ of the *missMethyl* R package^36^ to address probe number and multi-gene bias specific to methylation data from arrays. A list of DMPs and all CpGs tested were used as input, and both Gene Ontology (GO) and KEGG pathway collections were tested. We used a Benjamini-Hochberg adjusted p-value (FDR_B&H_) threshold of 0.05 to assess the statistical significance of enriched gene sets.

### Methylation age analysis

Methylation age is the prediction of biological age from methylation levels at a subset of CpGs (clock CpGs). PhenoAge^37^ and GrimAge^38^ are the most accurate and widely used methylation age algorithms, and have been associated with increased risk of various morbidities and mortality^37–39^.

We estimated methylation age using the PhenoAge^37^ algorithm with the methyAge function of the *ENmix* R package^40^. GrimAge was calculated with the online calculator at https://dnamage.genetics.ucla.edu/. MAA was defined as the residual term from regressing chronological age on methylation age estimates. For each algorithm, Shapiro-Wilk normality tests were used to test the normality of the MAA distribution. To test if mean MAA was significantly different between groups a one tailed t-test was used for the PhenoAge algorithm, and a Mann-Whitney test for the GrimAge algorithm.

## Results

### Cohort descriptive statistics

This study included 192 females with RMS across four study sites. Participants were categorised as nulligravida (n=96) or parous (n=96) based on available pregnancy history data. For parous participants, the median time from last conception to blood collection was 16.66 years (range = 1.45 – 44.42 years, **Table 1**).

### Differential methylation analysis – whole blood

After methylation data pre-processing, approximately 747,000 (86%) of 867,000 probes remained for differential methylation analysis (**Supplementary Fig. 2**). Batch effect analysis identified Plate, Sentrix ID and Sentrix Position as significant sources of technical variation (p < 0.01), which were corrected and reduced to negligible effects using the *Combat* algorithm^19^ (**Supplementary Fig. 2**).

Whole-blood methylation analysis revealed 903 differentially methylated positions (DMPs) surpassing genome-wide thresholds (FDR < 0.05 and Δ_meth_ > 1%, **Table 2** shows the top 10 DMPs by effect size (full list is available in **Supplementary Table 1**. DMPs mapped to 585 genes and 318 unannotated genomic locations. Of the 903 DMPs, 365 (40%) were hypomethylated and 528 (60%) were hypermethylated in the parous group relative to the nulligravida group (**Supplementary Fig. 3**). Δ_meth_ ranged from -13.28% to 16.10%. CpG islands are associated with gene promoter regions, in which methylation is likely to impact gene transcription. Only 106 (11.7%) of DMPs were in islands, with 173 (19.2%) in shores, 64 (7.1%) in shelves and the majority in open sea regions (560, 62.0%). Of 903 DMPs, five overlapped with the 10,592 DMPs identified by Mehta et al. (2019, **Supplementary Table 2**).

**Table 2.** Top 10 differentially methylated positions (DMPs) by effect size (Δ_meth_)

| CpG | CHR | MAPINFO | $\Delta_{\text{meth}}$ | FDR | Gene | Feature | CGI |
| --- | --- | --- | --- | --- | --- | --- | --- |
| cg12036633 | 15 | 63758958 | −0.161 | 0.036 |  | IGR | opensea |
| cg02122327 | 13 | 50194322 | 0.133 | 0.028 |  | IGR | opensea |
| cg03885684 | 2 | 120770471 | 0.108 | 0.014 | EPB41L5 | TSS200 | island |
| cg24000535 | 14 | 91110600 | −0.083 | 0.037 | LOC101928909 | Body | opensea |
| cg08166072 | 2 | 46213920 | 0.080 | 0.017 | PRKCE | Body | opensea |
| cg14248704 | 5 | 151470842 | 0.074 | 0.014 | CTB-12O2.1 | Body | opensea |
| cg10140164 | 9 | 75597328 | −0.072 | 0.029 |  | IGR | opensea |
| cg07723864 | 13 | 25670042 | −0.069 | 0.029 | PABPC3 | TSS1500 | island |
| cg06809965 | 14 | 70070333 | −0.063 | 0.024 |  | IGR | opensea |
| cg19938535 | 6 | 25341389 | 0.063 | 0.047 | LRRC16A | Body | opensea |

*Abbreviations: Chr = Chromosome, bp = base pair, CpG = cytosine-phosphate-guanine, CGI = CpG island, max = maximum, IGR = intergenic region, TSS200 = transcript start site (up to 200bp 5' of 5'UTR) promoter region, Body = gene body, TSS1500 = Transcript start site (up to 1500bp 5' of 5'UTR) promoter region*

No differentially methylated regions (DMRs) were identified using the *DMRcate* algorithm at an FDR threshold of 0.05. Therefore, we identified DMRs from our DMP list, defining a DMR as a region containing at least three DMPs with the same effect direction and FDR < 0.01, within 1000bp of the adjacent DMP/s^21^. Using this definition, we identified five DMRs on Chromosomes 7, 15, 17, 18 and 21 (**Table 3**). However, only DMR^Chr7^ (Δ_max =_ 0.029, FDR = 0.021, Supplementary **Fig. 4a**) and DMR^Chr15^ (Δ_max =_ 0.049, FDR = 0.015, Supplementary **Fig. 4b**) remained after mQTL analysis (see below). DMR^Chr7^ mapped to CRYGN, while DMR^Chr15^ mapped to an intergenic region. Both DMRs were hypermethylated in the parous group, relative to the nulligravida group.

**Table 3.** Differentially methylated region (DMR)

| Chr | Start (bp) | End (bp) | Width | n<br>CpGs | $\Delta_{\max}^*$ | $\Delta_{\text{mean}}^*$ | Gene | Feature | CGI | CpG Names |
| --- | --- | --- | --- | --- | --- | --- | --- | --- | --- | --- |
| 7 | 151137882 | 151138295 | 413 | 3 | 0.029 | 0.023 | CRYGN | TSS1500 | Shore | cg23666844,<br>cg16077872,<br>cg17362899 |
| 15 | 67228722 | 67228986 | 264 | 3 | 0.049 | 0.039 |  | IGR | Open<br>Sea | cg26795333,<br>cg20560283,<br>cg17174814 |
| 17 | 4803684 | 4804357 | 673 | 3 | -<br>0.045 | -0.042 | CHRNE | Body | Island | cg22349396,<br>cg06444025,<br>cg01726265 |
| 18 | 72152075 | 72152314 | 239 | 3 | 0.046 | 0.033 |  | IGR | Open<br>sea | cg23973972,<br>cg15682262,<br>cg27477494 |
| 21 | 44782331 | 44782497 | 166 | 4 | 0.022 | 0.018 | LINC01679 | TSS200,<br>IGR | Island | cg17577862,<br>cg02260098,<br>cg25191041,<br>cg14081667 |

\*Change from nulligravida to parous groups
*Abbreviations: Chr = chromosome, bp = base pair, CpG = cytosine-phosphate-guanine, CGI = CpG island, max = maximum, CRYGN =crystallin gamma N, CHRNE = cholinergic receptor nicotinic epsilon subunit, LINC01679 = long intergenic non-coding RNA 1679, TSS1500 = transcript start site (up to 1500bp 5' of 5'UTR) promoter region , TSS200 = transcript start site (up to 200bp 5' of 5'UTR) promoter region, IGR = intergenic region*

### Differential methylation analysis – immune cell specific

Differential methylation analysis of whole blood may not be sensitive to cell specific DMPs associated with outcome. Therefore, we estimated and compared the proportion of immune cell types between groups. There were no significant differences in immune cell type proportions between nulligravida and parous women (data not shown), and therefore we did not need to adjust our whole blood analysis for this variable. Statistical deconvolution revealed four CD4+ **(Table 4)** and eight CD8+ T cell specific DMPs (**Table 4**). All CD4+ T cell DMPs were hypermethylated in the parous group compared to the nulligravida group, and only one DMP mapped to a gene (cg14172633, *HMCN1*). In CD8+ T cells, three DMPs were hypermethylated and five were hypomethylated in the parous group. The DMP cg25577322 had the largest effect size (estimate = -8.32, SE = 1.45) and mapped to *AHR*. Seven of the eight DMPs mapped to a gene, and two DMPs mapped to *OR2L13* (cg08944170 and cg20507276).

**Table 4.** Cell specific differentially methylated positions (csDMPs)

| **CpG** | **Est.** | **SE** | **p** | **Nulligravida**<br>**mean\*** | **Parous**<br>**mean\*** | **Direction**<br>**of**<br>**effect\*\*** | **Chr** | **bp** | **Gene** | **Feature** |
| --- | --- | --- | --- | --- | --- | --- | --- | --- | --- | --- |
| *CD4+ T cells* |  |  |  |  |  |  |  |  |  |  |
| cg14172633 | -3.64 | 0.62 | 1.7x10<sup>-8</sup> | -3.10 | -2.84 | Hyper. | 1 | 185703557 | *HMCN1* | TSS200 |
| cg15145296 | -3.23 | 0.56 | 3.6x10<sup>-8</sup> | -4.24 | -3.98 | Hyper. | 3 | 125709740 |  |  |
| cg06032337 | -3.17 | 0.56 | 5.1x10<sup>-8</sup> | -2.82 | -2.71 | Hyper. | 6 | 29648468 |  |  |
| cg06818823 | -5.88 | 1.04 | 5.8x10<sup>-8</sup> | -6.55 | -5.90 | Hyper. | 6 | 46459236 |  |  |
| *CD8+ T cells* |  |  |  |  |  |  |  |  |  |  |
| cg01858500 | -4.27 | 0.71 | 1.1x10<sup>-8</sup> | -3.57 | -3.52 | Hyper. | 17 | 68202566 |  |  |
| cg08944170 | -2.78 | 0.48 | 2.3x10<sup>-8</sup> | -3.38 | -3.60 | Hypo. | 1 | 248100614 | *OR2L13* | 1stExon |
| cg25577322 | -8.32 | 1.45 | 4.0x10<sup>-8</sup> | -6.33 | -6.94 | Hypo. | 7 | 17338213 | *AHR* | TSS200 |
| cg16402757 | -2.14 | 0.38 | 4.6x10<sup>-8</sup> | -2.14 | -2.05 | Hyper | 10 | 35311004 | *CUL2* | Body |
| cg03495768 | -3.05 | 0.54 | 6.1x10<sup>-8</sup> | -2.98 | -3.07 | Hypo. | 13 | 100637113 | *ZIC2* | Body |
| cg04798314 | -2.71 | 0.48 | 6.7x10<sup>-8</sup> | -2.35 | -2.56 | Hypo. | 1 | 246668601 | *SMYD3* | Body |
| cg11738485 | -2.42 | 0.43 | 6.8x10<sup>-8</sup> | -2.79 | -2.51 | Hyper | 19 | 12877000 | *HOOK2* | Body |
| cg20507276 | -2.68 | 0.48 | 8.6x10<sup>-8</sup> | -3.40 | -3.55 | Hypo. | 1 | 248100600 | *OR2L13* | 1stExon |

\*Mean M-values reported as M-values used in cell-specific statistical analyses
\*\*Nulligravida to parous
Abbreviations: CpG = cytosine-phosphate-guanine, Est = estimate (from linear regression), SE = standard error, Hyper. = hypermethylated, Chr = chromosome, bp = base pair, TSS200 = transcript start site (up to 200bp 5' of 5'UTR) promoter region

### Sensitivity analysis

Sensitivity analyses revealed no major effects of symptom duration, ARR, cell type proportion estimates (B cells, CD4+ cells, CD8+ cells, NK cells, monocytes, and granulocytes), methylation age acceleration (PhenoAge and GrimAge), treatment at blood collection (yes or no) or smoking status at blood collection (ever or never) on differential methylation in this cohort, as demonstrated by the lack of association between CpGs and the covariates tested (data not shown). Therefore, these covariates were not included in the differential methylation analyses so as not to unnecessarily burden the model and reduce statistical power. One CpG (cg03708250) showed suggestive association with age at blood collection (FDR = 0.042) but was not identified as a DMP. Of the 2,622-smoking associated CpGs from Joehanes (2016) and 903 DMPs identified in this study, 25 overlapped and were removed prior to downstream analyses to avoid confounding (**Supplementary Table 3**).

### Methylation quantitative trait loci (mQTL) analyses

After quality control and filtering, 183 patients remained for mQTL analysis. SNVs located within 5kb up/downstream of each DMR were identified for LD. DMR^Chr7^ contained five independent SNVs (**Supplementary Table 4**). Methylation at cg23666844, cg16077872 and cg17362899 was associated with genotype at 7:151133104:G:C, 7:151135503:C:T, 7:151137301:G:C and 7:151140431:T:C **(Supplementary Table 5)**. After accounting for genotype at these SNVs, methylation at cg23666844, cg16077872 and cg17362899 remained associated with parity despite the presence of mQTLs (**Supplementary Table 5**).

**Table 5.** Top ten CpGs associated with parity as selected by the elastic net model.

| **CpG** | **Importance** | **Chr** | **Position (bp)** | **Gene** | **Feature** |
| --- | --- | --- | --- | --- | --- |
| **cg08186508** | 100.00 | 14 | 68067006 | *PIGH* | 5'UTR |
| **cg26506013** | 99.95 | 5 | 133984553 | *SEC24A* | 1stExon |
| **cg23841819** | 99.92 | 1 | 204970383 | *NFASC* | Body |
| **cg25485991** | 99.86 | 17 | 8066461 | *VAMP2* | TSS200 |
| **cg23367339** | 99.74 | 17 | 36622717 | *ARHGAP23* | Body |
| **cg17070338** | 99.69 | 13 | 111268441 | *CARKD* | Body |
| **cg07360021** | 99.49 | 6 | 151186904 | *MTHFD1L* | 1stExon |
| **cg11918372** | 99.14 | 2 | 48132755 | *FBXO11* | 5'UTR |
| **cg27573735** | 99.04 | 3 | 82857144 |  |  |
| **cg12835012** | 98.88 | 4 | 183795785 |  |  |

*Abbreviations: Chr = Chromosome, bp = base pairs*

DMR^Chr15^ contained five independent SNVs (**Supplementary Table 4**). Methylation at cg26795333 was associated with genotype at all SNVs. Methylation at cg20560283 and cg17174814 was associated with genotype at 15:67224485:A:G, 15:67224701:G:C, 15:67224979:C:G and 15:67228085:C:T **(Supplementary Table 5)**. After accounting for genotype at these SNVs, methylation at cg26795333, cg20560283 and cg17174814 remained associated with parity despite the presence of mQTLs (**Supplementary Table 5**).

DMR^Chr17^ contained four independent SNVs (**Supplementary Table 4**). Methylation at cg22349396 was associated with genotype at 17:67225730:G:C, 17:67226643:C:T and 17:67227383:A:T. Methylation at cg06444025 and cg01726265 was not significantly associated with genotype at any SNV (data not shown). However, methylation at all CpGs was no longer associated with parity after accounting for genotype at these four SNVs (**Supplementary Table 5**).

DMR^Chr18^ contained six independent SNVs (**Supplementary Table 4**). Methylation at cg23973972 was associated with genotype at 18:67224072:A:C, 18:67226036:G:A, 18:67226036:G:A and 18:67227787:A:C **(Supplementary Table 5)**. Methylation at cg15682262 was associated with genotype at 18:67224072:A:C, 18:67226036:G:A, 18:67226036:G:A and 18:67227787:A:C. Methylation at cg27477494 was associated with genotype at 18:67226036:G:A, 18:67226036:G:A and 18:67227787:A:C. After accounting for genotype at these SNVs, methylation at cg23973972, cg15682262 and cg27477494 was no longer associated with parity (**Supplementary Table 5**).

DMR^Chr21^ contained three independent SNVs: 21:44782007:C:T, 21:44782634:C:A and 21:44782732:A:C (**Supplementary Table 4**). Methylation at cg17577862, cg02260098, cg25191041 and cg14081667 was not associated with genotype at any SNV (data not shown). After accounting for genotype at these SNVs, methylation at cg17577862 remained associated with parity, however, methylation at cg02260098, cg25191041, cg14081667 was no longer associated with parity (**Supplementary Table 5**).

After accounting for genotype at independent SNVs located within (plus 5kb up/downstream) of each DMR, only DMR^Chr7^ and DMR^Chr15^ contained enough DMPs (≥ 3) to be considered DMRs.

### Multi-factor feature selection

Using elastic net regression, we identified a panel of CpGs conferring a conjoint association. We determined the optimal alpha (0.1) and lambda (0.02) values for our data using a cross validation approach. With an alpha value closer to zero than one, our elastic net regression resembled a lasso regression more closely than a ridge regression.

Using these model parameters, our elastic net regression model selected 1556 CpGs associated with parity (top 10 shown in **Table 5**, full list in **Supplementary Table 6** in our training dataset (n=134, 70% of cohort). Of these, 322 CpGs (34.5%) were also identified as DMPs in our primary analysis. The most important CpG in the model, cg08186508 (variable importance = 100), is located on Chromosome 14 and maps to the *PIGH* gene. However, cg08186508 was not identified as a DMP in our primary analysis suggesting its effect is in correlation with other CpGs.

### Gene-set enrichment analysis (GSEA)

We conducted GSEA on all differentially methylated genes identified in the primary methylation analysis and elastic net regression to elucidate potentially small but cumulative effects of parity on methylation patterns (**Supplementary Table 7**). 609 of 903 DMPs (67%), and 1208 of the 1556 CpGs (78%) identified in the elastic net regression, mapped to a gene. In total, 1318 unique genes were used for GSEA.

ToppGene revealed that differential methylation, regardless of direction of effect, was primarily enriched in biological processes (**Figure 1a**) and cellular compartments (**Figure 1b**) related to neuroplasticity, including neurogenesis (n_genes_ = 178, FDR_B&H_ = 2.77×10^−5^), neuron projection morphogenesis (n_genes_ = 87, FDR_B&H_ = 2.77×10^−5^) and neuron projection (n_genes_ = 175, FDR_B&H_ = 6.96×10^−10^). Furthermore, the top enriched molecular functions related to ion transport including cell adhesion molecule binding (n_genes_ = 70, FDR_B&H_ = 2.06×10^−4^) and GTPase regulator activity (n_genes_ = 61, FDR_B&H_ = 3.29×10^−5^, **Figure 1c**).

**Figure 1.**
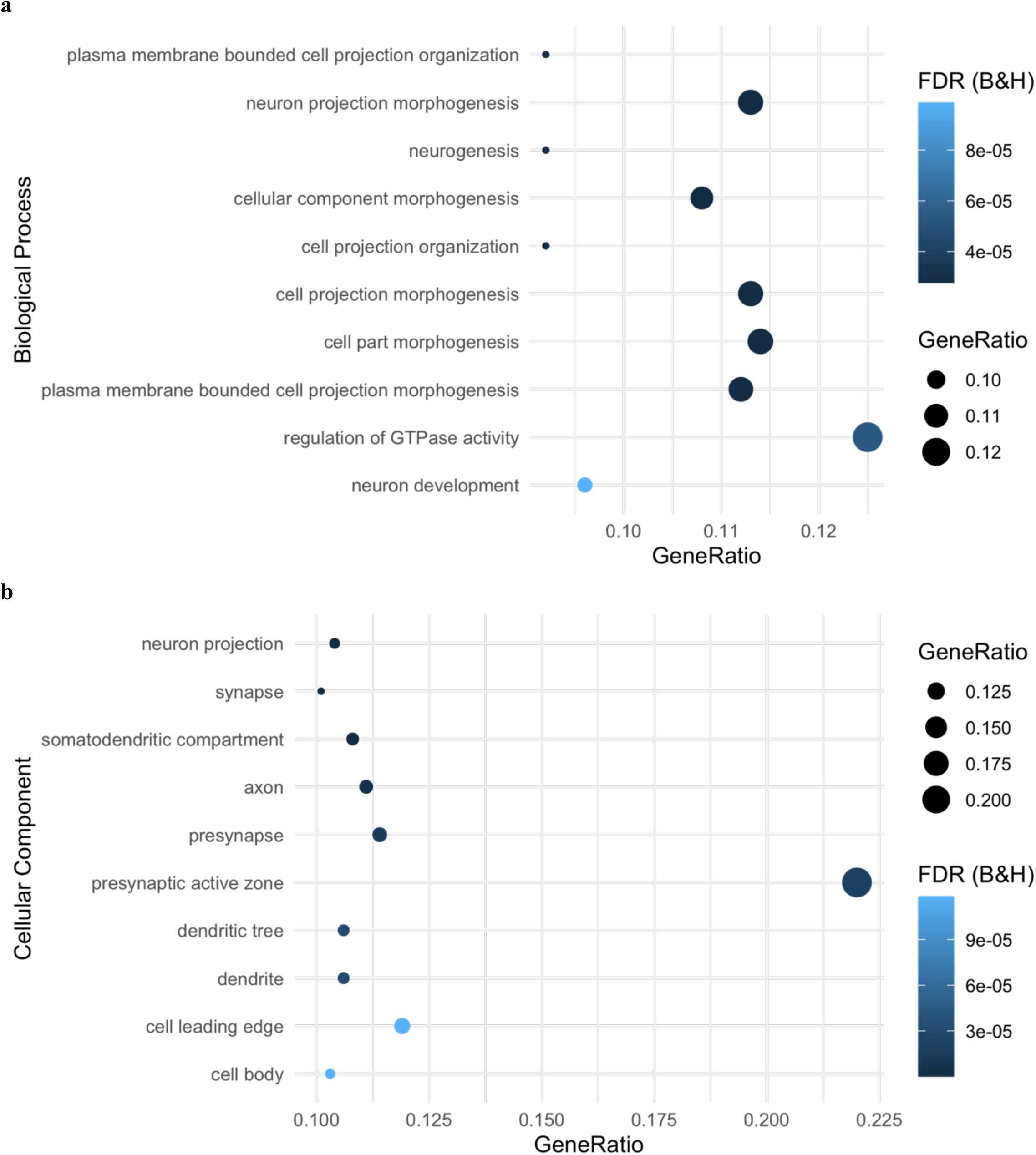
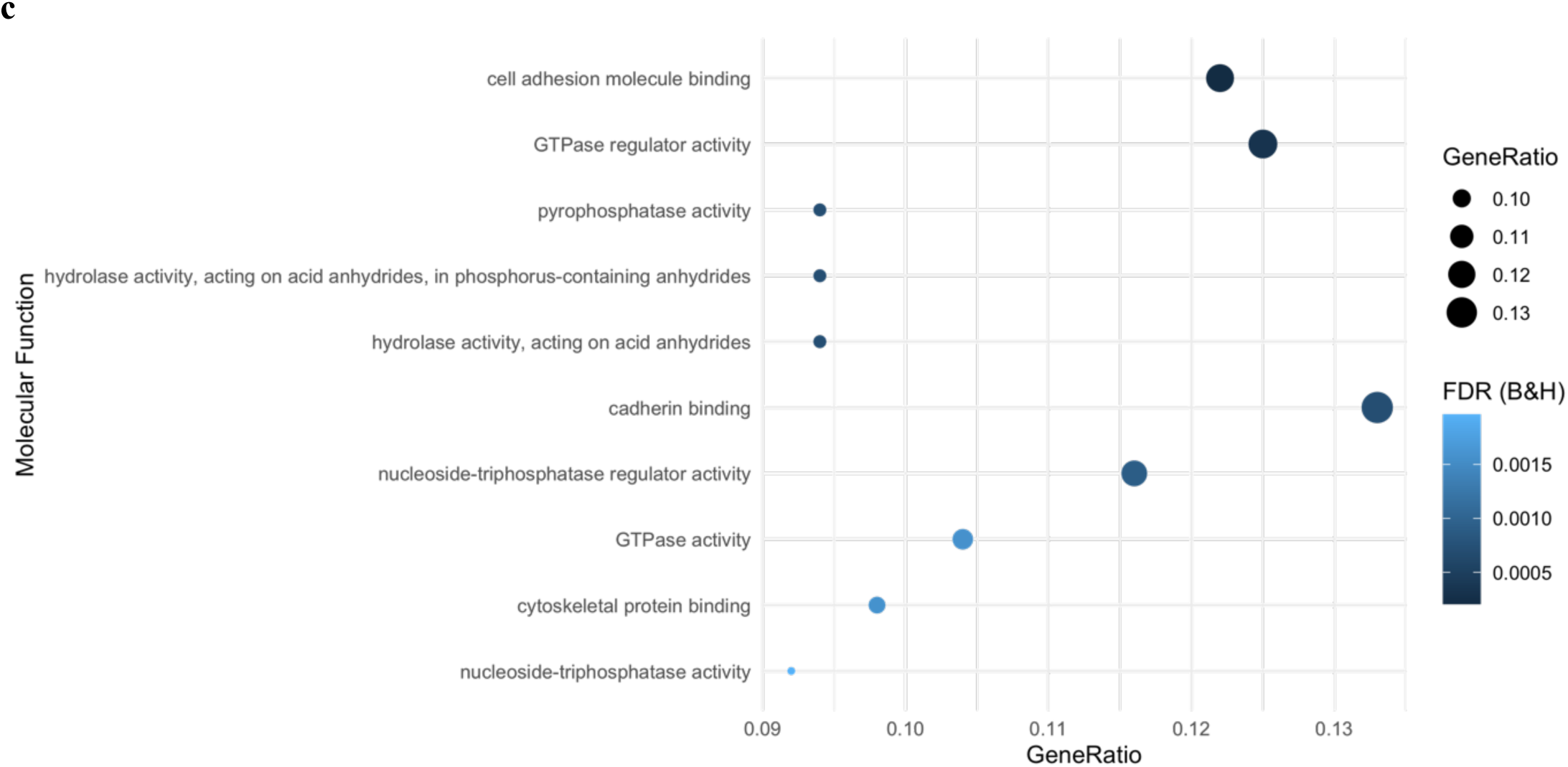
Gene Set Enrichment Analysis (GSEA) using ToppGene API. Input data were genes identified in both the differential methylation analysis and elastic net regression (n=1318). The **a)** ten most significantly enriched biological processes, **b)** ten most significantly enriched cellular compartments, **c)** ten most significantly enriched molecular functions. Gene ratio is the ratio of the number of genes in the query list and the hit count for that gene set in the genome.

Hypomethylated genes (n=576) drove the enrichment of neuron development and growth biological processes and cellular compartments. Alternatively, hypermethylated genes (n=775) drove the enrichment of signal transduction biological processes and molecular functions.

There were no enriched gene ontology terms using GOmeth with an FDR threshold of 0.05. This suggests that our ToppGene findings could be a result of probe number or multi-gene bias. However, we used GSEA as an exploratory analysis to generate hypotheses about the mechanism in which pregnancy impacts MS clinical outcomes and have therefore interpreted the results with caution.

### Methylation age analysis

Methylation Age Acceleration (MAA) measures the disparity between chronological and biological age as estimated using methylation age algorithms, and can provide insight into an individual’s health and lifespan^39,37,38^. As groups were *a priori* matched by age, there were no significant differences in chronological age between groups (**Table 1)**.

The correlation between chronological age and methylation age using the PhenoAge and GrimAge algorithms were 0.77 and 0.91, respectively. We did not find any evidence for differences in methylation age between groups using the GrimAge algorithm (p = 0.854). However, we did find significant differences in methylation age between groups using the PhenoAge algorithm (p = 0.034, **Supplementary Fig. 5)**.

MAA was calculated as the residual term from regressing chronological age on methylation age. Residual terms were normally distributed for the PhenoAge (p = 0.551) algorithm, but not the GrimAge algorithm (p = 3.52×10^−05^). There were significant differences in MAA between nulligravida and parous groups using both the PhenoAge (mean difference = 2.27 years, p = 0.001) and GrimAge algorithms (mean difference = 1.44 years, p=0. 0.005, **Figure 2**).

**Figure 2.**
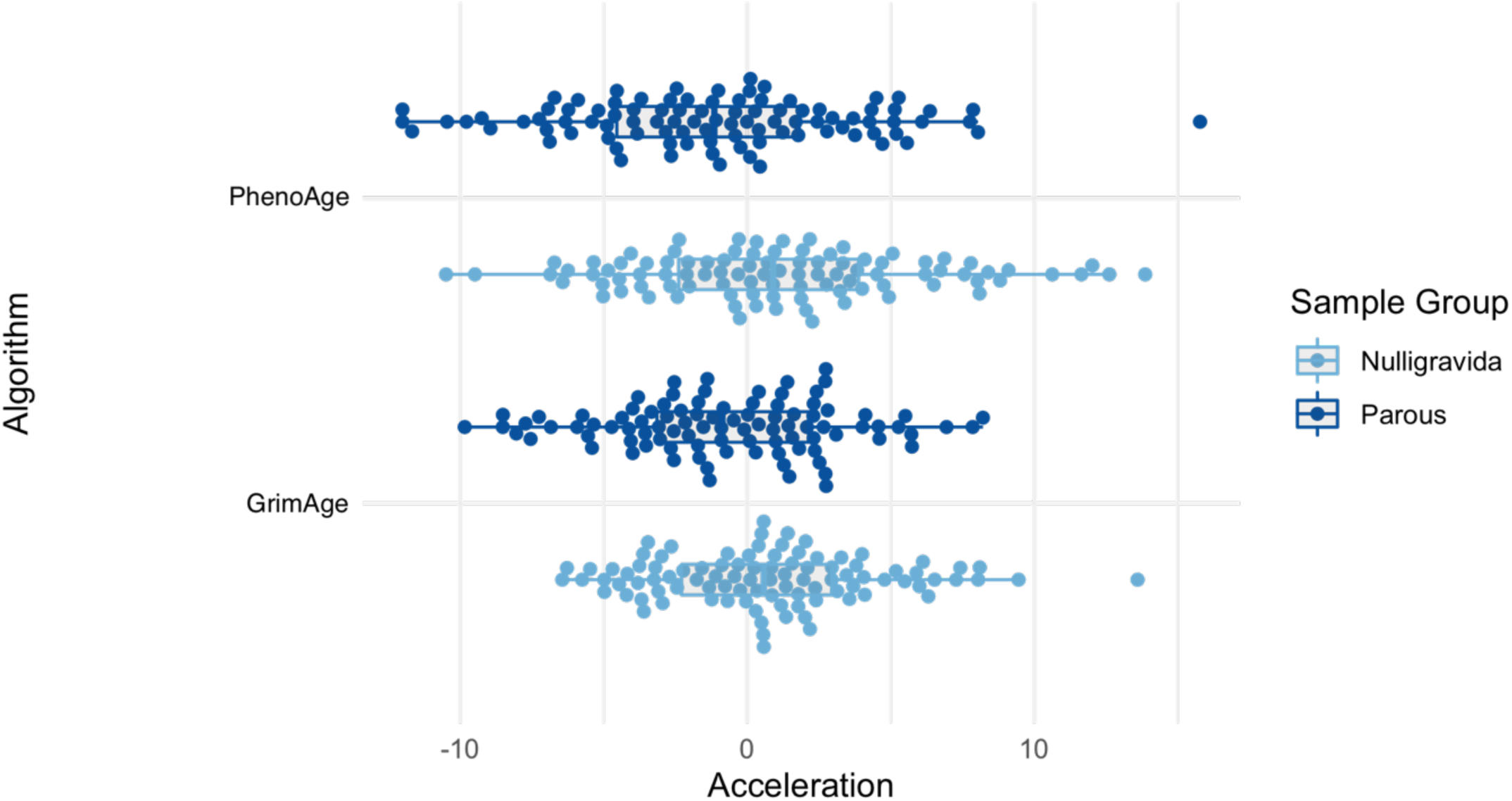
Methylation age acceleration (MAA) by sample group using the PhenoAge and GrimAge algorithms. There are significant differences in MAA between groups using the PhenoAge and GrimAge algorithms. PhenoAge: nulligravida mean = 1.14 (SE = 0.502), parous mean = -1.14 (SE = 0.504), mean difference = 2.27 years, p = 0.001. GrimAge: nulligravida mean = 0.720 (SE = 0.749), parous mean = -0.720 (SE = 856), mean difference = 1.44 years, p=0. 0.005.

## Discussion

Studies have demonstrated an association between pregnancy and reduced disability accumulation in women with MS (wwMS)^2^, lasting for up to ten years post pregnancy^4^. Recent studies have identified associations between birth history and methylation patterns in health^41–43^ and MS^8^, as well as negative associations between birth history and methylation age acceleration in women without MS^6^. No methylome-wide studies to date have examined associations between methylation patterns and parity, or methylation age acceleration in wwMS.

Our primary EWAS of whole blood methylation differences between nulligravida and parous wwMS identified 903 differentially methylated positions (DMPs) across autosomes. Of the DMPs identified in our study, five overlapped with those previously identified by Mehta and colleagues (2019)^8^. This is reasonably explained by differences in cohort size and study design, where Mehta et al. (2019) sought to identify DMPs in genes that were identified *a priori*^8^, compared to our genome-wide approach. Moreover, they included women with a history of pregnancy, compared to our study which included only women with a history of birth. The overlapping DMPs mapped to *PRIC285, GRTP1, SIM2 and CCDC90B*, and one intergenic region. *PRIC285* is a nuclear transcriptional co-activator and an interferon effector gene that is integral to antiviral immune responses^44^. *GRTP1*, is a GTPase activator associated with platelet counts^45^. Notably, *SIM2* is a transcription factor and master regulator of central nervous system development and neurogenesis^46^, and a link between neurogenesis, neuronal reserve and MS outcomes is frequently hypothesised^47^. Further, *CCDC90B* is a protein coding gene which regulates mitochondrial calcium ion concentrations affecting ATP production^45^. A link between mitochondrial dysfunction in neurons and MS outcomes has recently been identified^48^. While we identified a different direction of effect at *CCDC90B* between our study and Mehta et al. (2019), this may be due differences in sample size or timing of sample collection. In this study, we confirm that genes related to these processes are differentially methylated between nulligravida and parous wwMS for up to 44.4 years post-pregnancy. Single nucleotide variants associated with mitochondrial and CNS function were recently shown to associate with MS severity outcomes^49^. Our study demonstrates a putative mechanism by which pregnancy may impact long-term legacy effects on outcomes in wwMS.

In addition to 903 DMPs, we identified two differentially methylation regions (DMRs) on Chromosome 7 and 15. DMR^Chr7^ contains three DMPs in the transcript start site (up to 1500bp 5’ of 5’UTR) promoter region of *CRYGN* which encodes crystallin gamma N, a structural protein in eye lenses. The literature on *CRYGN* is limited, and it has not previously been linked to pregnancy or MS. Our finding requires validation in an independent cohort and replicated DMP/DMR signals would provide a strong rationale for in vitro functional studies of gene and protein expression control mediated by the DMR.

Using statistical deconvolution, we identified four CD4+ T cell specific DMPs (csDMPs). CD4+ T cells are central to immune regulation and tolerance, and have been strongly linked to both MS^50^ and pregnancy^51^. Multiple studies have reported changes in the epigenetic patterns of CD4+ T cells during pregnancy in wwMS^7,52,53^. The only gene-associated DMP was at the transcription start site for *hemicentin 1* (*HMCN1)*, a member of the immunoglobulin superfamily. However, to date *HMCN1* has not been associated with differential methylation in healthy populations or wwMS during pregnancy^52^. Nor is there literature linking *HMCN1* to MS risk. Therefore, this finding, together with the association of differential methylation of *CRYGN* is highly novel and requires independent validation.

We also identified eight CD8+ T cell csDMPs that map to six genes and one intergenic region. The involvement of CD8+ T cells in MS pathophysiology is well established^50^. During pregnancy CD8+ T cells are critical for maternal–fetal tolerance and protection against viruses^54^. The functions and diseases associated with the CD8+ T cell csDMPs identified in this study suggest they are markers of pregnancy outcomes, rather than genes implicated in the modulation of MS outcomes due to pregnancy (e.g., *OR2L1, HOOK2 and CUL2)*. Most notably, Aryl Hydrocarbon Receptor (*AHR*, cg25577322) is upregulated in decidual natural killer cells in women with recurrent spontaneous abortion and was hypomethylated in parous women in our study^55^. Here, we excluded pregnancies ending in miscarriage or termination to prevent identifying epigenetic biomarkers of miscarriage or termination. While it is possible that this signal was driven by unreported terminations and/or unknown miscarriages, it was identified in peripheral CD8+ T cells only (not whole blood) and is therefore unlikely to be a marker of recurrent spontaneous abortion in this cohort.

Furthermore, multiple studies have recently correlated *AHR* agonist activity with MS subtype and prognosis^56,57^. In these studies, *AHR* agonist activity increase was associated with relapse in CIS and RRMS^56^. A decrease in *AHR* agonist activity was associated with RRMS remission^56^ and progressive MS,^57^ thus implicating *AHR* in neuroinflammatory processes. In our study, *AHR* was hypomethylated. Hypomethylation is often, but not always associated with upregulation of gene expression. Unfortunately, we did not assess gene expression. However, this provides a plausible mechanism by which pregnancy could modulate disease outcomes and warrants further investigation.

We conducted GSEA on 1318 differentially methylated genes identified in the primary analysis and elastic net regression to generate hypotheses about the functional role of these genes in long-term MS outcomes. Hypomethylated genes in parous wwMS were enriched in neuron development and growth biological processes and cellular compartments, and hypermethylated genes were enriched in signal transduction biological processes and molecular functions. Mehta et al. (2019) similarly found enrichment of neuronal pathways including axon guidance in their study of differentially expressed genes between nulliparous and parous wwMS^8^. While the majority of DMPs (95.9%) in our primary differential methylation analysis had small effect sizes, the strength of our penalised regression approach is the ability to reveal small, correlated relationships between features. Taken together, these findings suggest that methylation impacts neuraxonal maintenance and neurite growth in parous women in a small but cumulative manner, up to 44.4 years after pregnancy. These findings are consistent with reports that the brains of women who have children undergo pronounced morphological changes as a result of pregnancy^58^.

Ours is the first study to report a reduction in MAA in parous wwMS, compared to age-matched nulligravida wwMS. We demonstrated that parous women have a reduced mean MAA of between 1.44 - 2.27 years depending on the algorithm employed. This shows that, as in health, parity is associated with a reduction in MAA in wwMS^6^. GrimAge is the newest algorithm with robust associations with morbidity and mortality^38^. Furthermore, PhenoAge acceleration is associated with an increased risk of physical functioning problems^59^. As a whole, our findings demonstrate slower biological aging in parous wwMS, and potentially a longer period of health and lifespan^59^.

Ours is the largest study to date investigating the association between genome-wide methylation and parity in women with relapse-onset MS. We identified hundreds of methylation changes associated with parity that may underlie long-term outcomes in wwMS. Cohort matching by age limited confounding and erroneous associations between methylation patterns and parity. We aimed to mitigate against confounding by disease severity by matching for ARMSS scores, therefore allowing us to study the relationship between methylation patterns and parity specifically. Whether these changes are specific to wwMS or a broader response to pregnancy remain to be confirmed in future studies. We were underpowered to adjust for a range of clinical and environmental factors potentially associated with methylation patterns, including number of births and DMT^60^. Study power also limited our ability to identify small cell type-specific effects beyond those identified in T cells. Therefore, our findings require validation in a larger, independent cohort of wwMS. As ours is a retrospective and cross-sectional study, we were not able to establish a causal link between pregnancy, methylation pattern changes, and long-term clinical outcomes in wwMS. We are currently undertaking a longitudinal and prospective study of methylation changes during and after pregnancy relative to a nulligravida baseline, to investigate temporal and causal relationships between pregnancy, methylation, and disease outcomes in wwMS. This could lead to the identification novel therapeutic targets.

## Conclusion

We investigated the association between whole blood and cell type specific genome-wide methylation patterns and parity in 192 women with relapse-onset MS. We identified small but potentially cumulative differences in whole-blood and T-cell methylation patterns in genes related to neural plasticity, offering a putative molecular mechanism driving the long-term effect of pregnancy on MS outcomes. We further identified reduced methylation age acceleration in parous wwMS, demonstrating slower biological aging compared to nulligravida wwMS. As methylation patterns can be cell type specific, our results suggest a potential ‘CNS signature’ of methylation in peripheral immune cells, as previously described in relation to MS progression^61^. This is the first genome-wide methylation study of parity in wwMS and therefore, validation studies are needed to confirm our findings.

## Data Availability

Access requests for methylation data with scientifically sound proposals can be made in writing to Dr. Vilija Jokubaitis and Prof. Jeannette Lechner-Scott. Requests will be assessed and responded to within 4 weeks. If data access is approved, authorship should include the MSBase Genetics Consortium, with individual authors listed as appropriate.

## Data availability

Supplemental files contain data supporting the conclusions in this article. Data access requests with scientifically sound proposals can be made in writing to Dr Vilija Jokubaitis. Clinical data from the MSBase Registry: To protect participant confidentiality, de-identified patient-level data sharing may be possible in principle but will require permissions/consent from each contributing data controller.

## Acknowledgements

We thank all the people with MS who participated in this research without whom this work would not be possible. The authors would like to acknowledge research nurses Ms Jo Baker, Ms Jodi Haartsen, Ms Sandra Williams, Ms Lisa Taylor for assisting with blood collection for this study, and Dr Louise Laverick, and Ms Malgorzata Krupa for research assistance.

## Author declarations

None.

## Author contributions

MPC conducted statistical analyses, drafted, and revised the manuscript and aided in study design and data interpretation. VM aided in sample collection, data interpretation and revised the manuscript. MS and TK aided in sample collection and revised the manuscript. HB and JLS aided in sample collection, study design, data interpretation and revised the manuscript. RS acquired funding, helped develop the concept, aided in data interpretation, and revised the manuscript. RL and VJ designed and conceptualised the study, acquired study funding, aided in sample collection and data interpretation, and drafted and revised the manuscript.

