## Supplementary Figures for "Birth history is associated with whole-blood and T-cell methylation patterns in relapse onset multiple sclerosis"

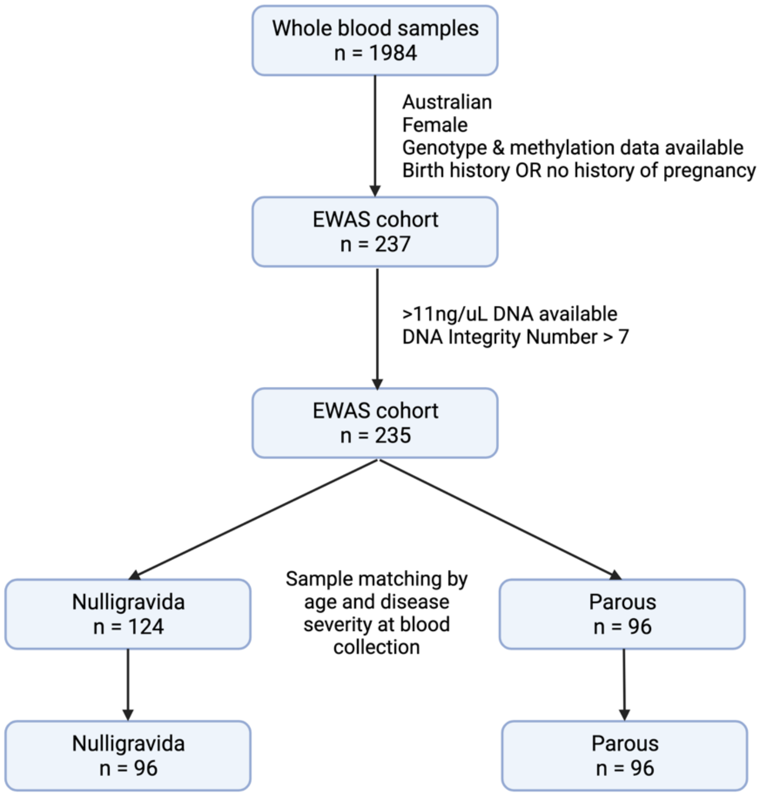


**Supplementary Figure 1. Participant inclusion flowchart.** Whole-blood samples were obtained from 1,984 participants. The cohort of 1,984 refined to 192 participants based on geographical location (Australia), sex (female), pregnancy history (nulligravida or parous), DNA quantity and quality, and age and disease severity at blood collection, as measured by Age Related Multiple Sclerosis Severity (ARMSS) scores.

*Created with BioRender.com*

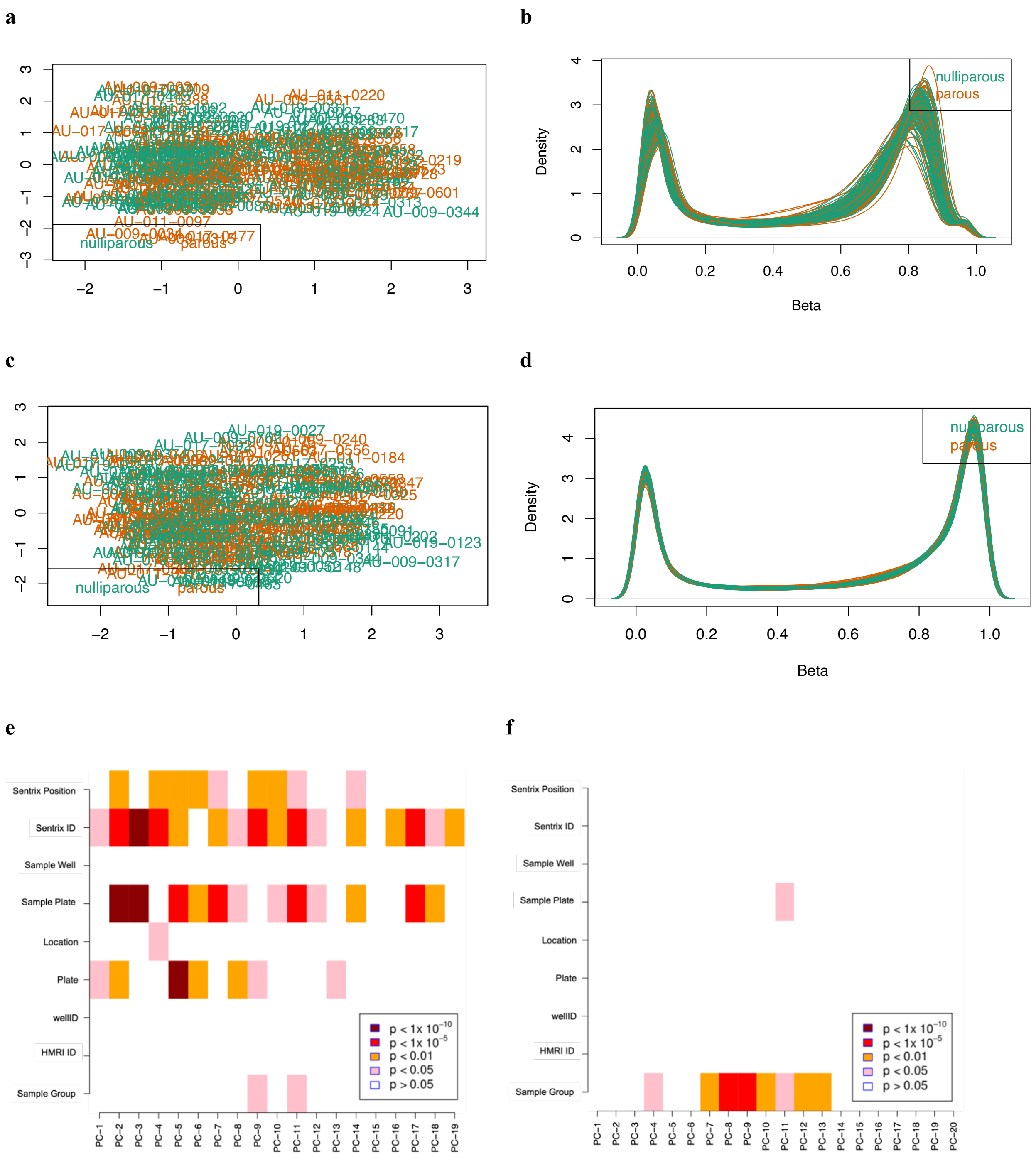
a
b
nulliparous
parous
Density
Beta
c
d
nulliparous
parous
Density
Beta
e
f
Sentrix Position
Sentrix ID
Sample Well
Sample Plate
Location
Plate
wellID
HMRI ID
Sample Group
p < 1x 10^-10
p < 1x 10^-5
p < 0.01
p < 0.05
p > 0.05

**Supplementary Figure 2. Quality control plots produced in ChAMP. a)** Multidimensional Scaling (MDS) Plot of methylation beta values for all samples, with the 1000 most variable positions displayed. There is no grouping of nulligravida or parous groups evident **b)** A density plot of raw methylation data prior to normalisation, plotting the beta value distribution for all samples. **c)** MDS plot after beta value normalisation. **d)** Density plot after beta value normalisation. Singular Value Decomposition (SVD) heatmaps **e)** before and **f)** after the removal of technical variation (batch effects) with ComBat. Once technical variation at Plate, Sentrix ID and Sentrix Position were removed with ComBat, Sample Group explained most of the variance in the data, as expected as the variable of interest.

*Abbreviations: MDS = Multidimensional Scaling*

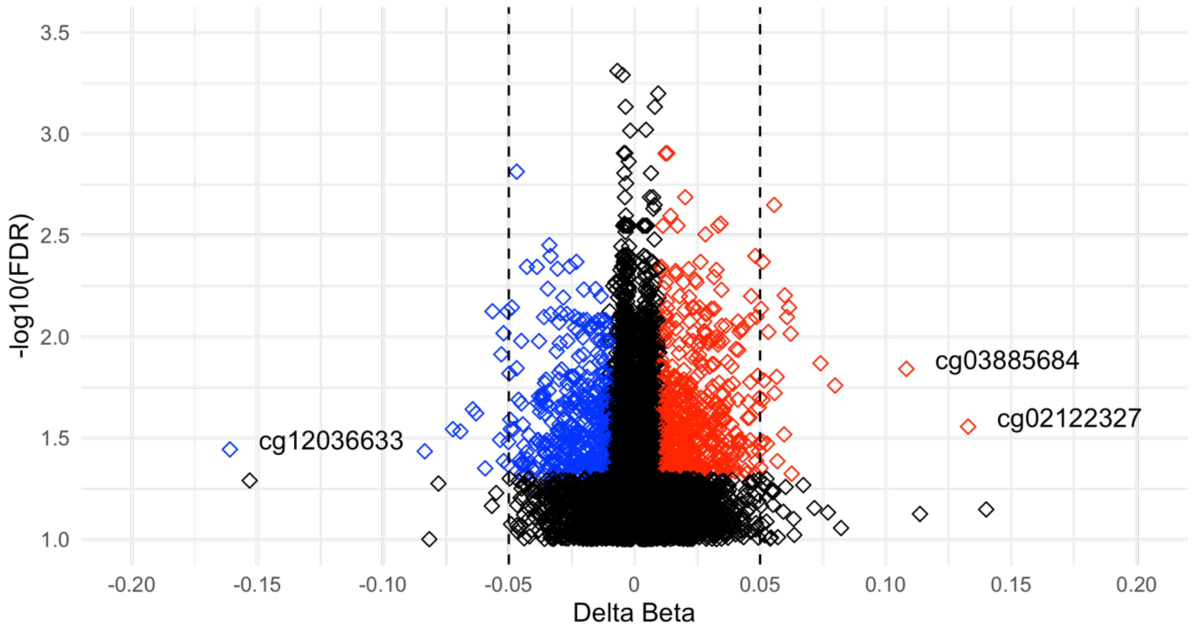


**Supplementary Figure 3. Differentially methylated positions (DMPs) between nulligravida and parous groups.** Of 903 DMPs, 365 (40%) were hypomethylated (blue) and 528 (60%) were hypermethylated (red) in the parous group. A delta beta (x-axis) of 0.1 is analogous to $\Delta_{meth}$ of 10% between groups. $\Delta_{meth}$ ranged from −13.28% to 16.10%. DMPs with $\Delta_{meth}$ above 10% are labelled.

*Abbreviations: FDR = false discovery rate*

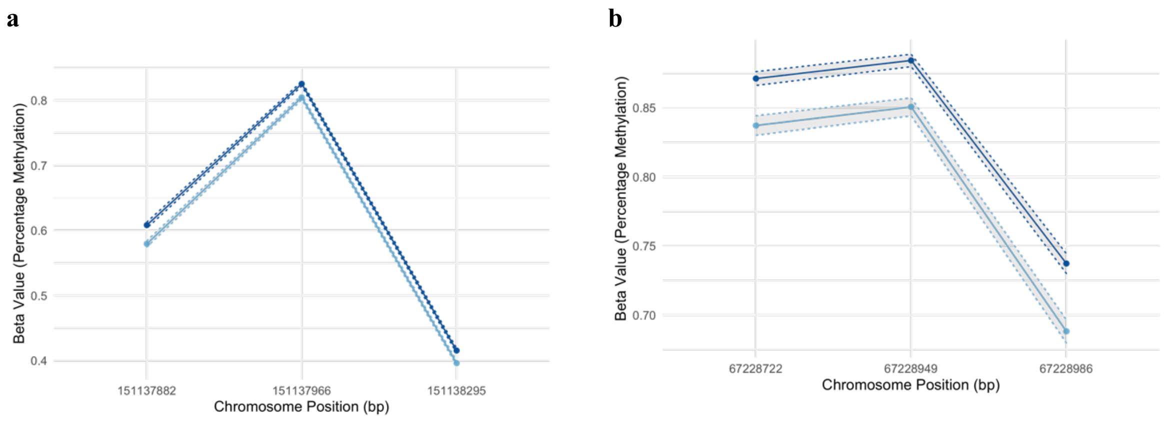


**Supplementary Figure 4. Differentially methylated regions (DMRs) between nulligravida and parous groups.** Mean methylation (β) values for nulligravida (light blue) and parous (dark blue) groups at each CpG in **a)** $DMR^{Chr7}$: Chr7:151137882-151138295, $\Delta_{max} = 0.029$, and **b)** $DMR^{Chr15}$: Chr15: 67228722-67228986, $\Delta_{max} = 0.049$. Grey shading shows standard error of the mean.

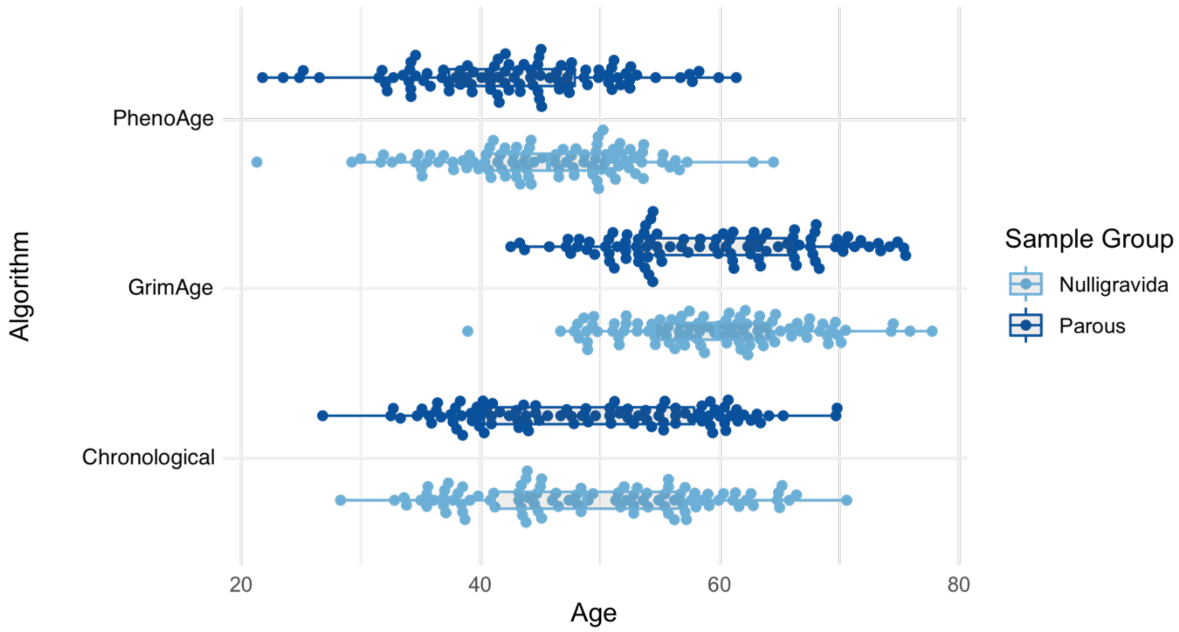


**Supplementary Figure 5. Methylation age estimates and chronological age by sample group.** Methylation age was estimated using the PhenoAge and GrimAge algorithms. Significant differences between groups were identified using the PhenoAge algorithm ($p = 0.034$). No significant differences between groups were identified using the GrimAge algorithm ($p = 0.854$).
